# Case Study of Nigeria’s experience scaling up chlorhexidine use in newborns, 2016-2021: analysis of implementation of actions to support scale-up and progress towards institutionalization

**DOI:** 10.1101/2025.10.22.25338605

**Authors:** Jim Ricca, Olayinka Badmus, Bose Adeniran, Femi James, Abdullahi Abdullahi Jatau, Edward Iyemi, Nosa Orobaton, Laura Ghiron, Young-Mi Kim, Diwakar Mohan, Regien Biesma

## Abstract

**Background:** Application of chlorhexidine (CHX) to the umbilical stump is an evidence-based intervention to prevent newborn infections. Successful pilot experiences led the Nigeria Federal Ministry of Health (FMoH) to launch a national, five-year scale-up strategy in 2016. In this case study, we analyze the implementation of national and state-level scale-up actions described and mandated in the national strategy and progress toward institutionalizing CHX within the routine health system using the WHO building blocks framework.

**Methods:** We conducted a mixed methods case study that consisted of interviews with key informants after the midline of the national strategy; focus groups of well-informed senior stakeholders at midline and endline of the strategy to assess the level of institutionalization achieved and the status of implementation of national-level scale-up actions described in the national strategy; and online surveys of state reproductive health coordinators (SRHCs) at midline and endline about implementation of state-level actions and the extent of international partner support.

**Results:** Institutionalization of CHX progressed steadily across all health system building blocks. Most rapid progress was in governance, followed by human resources and supply chain management, with slower progress on the health information system. The building block lagging most was financing, with continued dependence on external funding. Most but not all national-level actions were implemented as planned. There was wide variation in the implementation of state-level actions included in the national plan.

**Discussion and Conclusion:** Nigeria has made substantial progress in institutionalizing CHX across the health system building blocks. In terms of implementation of planned state-level scale-up actions, wide variation points to the need for more sustained and coordinated action, and the importance of actively managing this coordination. At national level, as Nigeria continues in the second phase of its scale-up efforts, the FMoH needs to strengthen coordination and financing.

## INTRODUCTION

Globally, in low- and middle-income countries (LMICs) overwhelming infection (sepsis) is one of the three leading causes of newborn death, along with prematurity and intrapartum-related complications [1]. Chlorhexidine digluconate 7.1% gel (CHX) is an over-the-counter broad-spectrum antiseptic. In 2007-8, several trials in LMICs established CHX’s effectiveness at preventing deaths from newborn sepsis when applied to the umbilical cord stump immediately after delivery and daily during the first week of life [2,3]. A meta-analysis of trials in several high mortality LMIC settings showed 23% reduction in all-cause mortality [4]. After these promising results, several technical agencies began promoting the use of CHX, including the global Chlorhexidine Working Group led by PATH [5]. In 2013, the World Health Organization (WHO) endorsed the use of CHX for infants born outside of health facilities in countries where the newborn mortality rate (NMR) is greater than 30 per 1,000 live births; there is a high proportion of home births; and there is prevalent use of harmful traditional substances [6]. WHO therefore included chlorhexidine digluconate 7.1% gel on its Essential Medicines List in 2013 [7]. Two later trials did not show positive results in lower mortality settings [8,9]. This contributed to WHO maintaining its recommendations for use of chlorhexidine in limited circumstances (i.e., not to recommend it for facility-based settings). However, given the potential complexity of communicating a strategy to scale up chlorhexidine in some settings and not others, several countries, including Nigeria, chose to encourage its use in all birth settings [10,11] (for Nigeria: personal communication with Director of Maternal and Child Health, Nigeria Federal Ministry of Health, June 2024).

Published frameworks and guidance on scaling up health innovations exist that are mainly based on programmatic experience [12,13,14,15]. Although the 2016 Nigeria national strategy (explained in more detail below) did not explicitly apply a particular framework, it applied many of the principles and common best practices outlined in this corpus of guidance documents. Among these frameworks, that of ExpandNet is the most explicit about the need not only for so-called ‘horizontal spread’ of the intervention, but also for institutionalization (what it terms ‘vertical scale up’), so that the new intervention is embedded in country systems and becomes a sustainable part of routine care [16]. The concept of institutionalization for a new intervention being scaled up has been described in more operational terms by researchers looking at the adoption and expansion of community-based child illness care in various countries (co-called Integrated Community Case Management, or iCCM). These authors defined institutionalization as “occur[ing] when the intervention becomes a routinely practiced and integral part of the conventional health system.” [17] This is the definition used for this study, and as those authors had, we looked at institutionalization (i.e., integration) of the CHX intervention within the Nigerian health system, as categorized by the World Health Organization’s health system building blocks [18].

The detailed nature of the national CHX scale-up strategy and the decentralization of the Nigerian health system set up the conditions for a natural experiment of sorts, offering an opportunity to gain insights from the differential levels of implementation of the various actions mandated in the national plan, across the various states, over time. It should be noted that there were independent national household health surveys at the baseline, midline, and endline of the national strategy. The 2016 Multiple Indicator Cluster Survey (MICS) [19] was done near the beginning of the CHX national scale-up strategy and confirmed near-zero baseline population coverage for CHX use, then another independent national household survey was done near midline of the national strategy period (2018 Demographic and Health Survey (DHS)) [20] showing a rapid rise to 11% population coverage and finally another independent national household survey (MICS 2021) [21] done near the end of the national strategy period showed that population coverage had plateaued. Analysis of this horizontal spread of CHX use is important background information, but it is not the focus of this study, which instead poses questions about implementation of the actions in the national CHX strategy and institutionalization of CHX in health systems:

- What progress has Nigeria made in implementation of scale-up actions envisaged in its national strategy?
- What progress has Nigeria made towards institutionalizing support for CHX in its health system building blocks (as described by the World Health Organization (WHO) [18]?

This study builds on a previously disseminated mixed methods case study of the scale-up process done in 2018-2019 that assessed progress and challenges of the national experience up to that point [22]

### Introduction and scale up of CHX use to prevent newborn sepsis in Nigeria

Nigeria has a complex and decentralized healthcare system and faces challenges in ensuring equitable access to quality healthcare services, especially in rural and underserved areas. In 2013, Nigeria’s neonatal mortality rate was estimated to be 38 per 1,000 live births, one of the highest in the world [23]. About 15% of neonatal deaths in 2016 were due to sepsis [24], many of which could be prevented through improving newborn umbilical cord care.

Nigerian public health program managers first learned about CHX as an evidence-based practice in 2012 during a conference of government and non-governmental stakeholders facilitated by the U.S. Agency for International Development (USAID)-funded Targeted States High Impact (TSHIP) project, which then supported the state ministries of health of the northern states of Sokoto and Bauchi to pilot implementation [25]. CHX was being introduced with the intention of displacing either dry-cord care or use of methylated spirits among those born in facilities, and the use of potentially harmful substances like toothpaste among those born in the community [26]. Sokoto state piloted CHX use mainly through community-based distribution and Bauchi state through public-health facilities. The Federal Ministry of Health (FMoH) also encouraged local production of CHX gel by private manufacturers to help keep product cost low, which resulted in several Nigerian pharmaceutical firms beginning production and distribution.

Positive results from the initial state-level experiences provided context-specific evidence that motivated the FMoH to promote national scale up of CHX use. In collaboration with various technical agency partners, and facilitated by Dalberg Consulting, in 2016 the FMoH developed and published a *National Strategy for Scale-up of Chlorhexidine in Nigeria 2016-2021* [26]. The strategy set a target of 52% coverage of CHX for all births in Nigeria by 2021, which the strategy document estimated would avert 55,000 neonatal deaths by 2021. It specified the use of 7.1% CHX gel in 25-gram tubes for once-daily application to the umbilical stump for seven days, starting on the first day of life, regardless of the location of birth. It identified organizations, individuals, and stakeholder groups to facilitate coordination of scale-up efforts across the multiple stakeholders to implement various actions needed to achieve the goals outlined in the national strategy, a summary of which is shown in Figure 1 [26]

**Figure 1:**
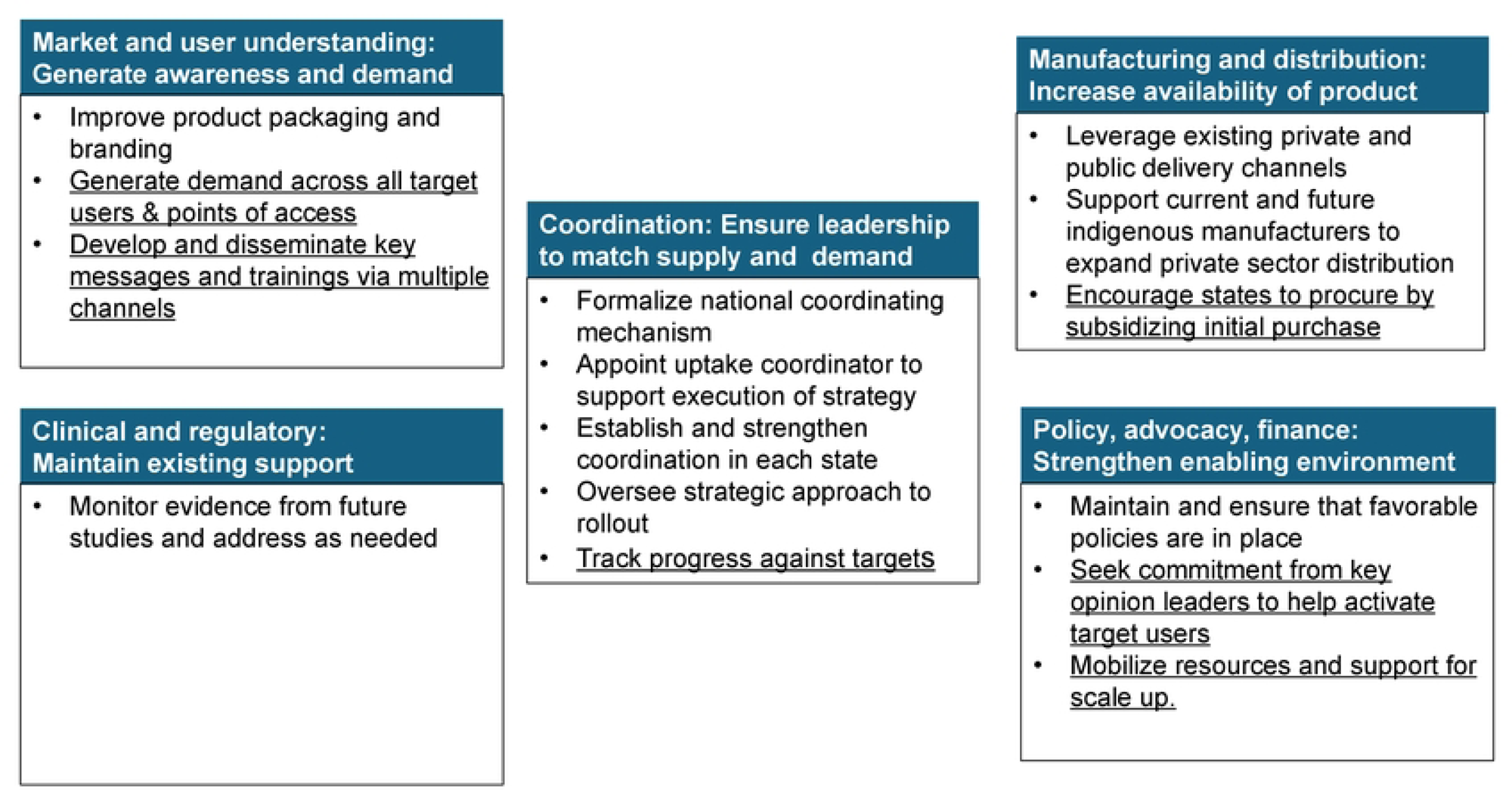
National CHX scale-up strategy and implementation plan (2016-2021) [26]. State level actions are <u>underlined</u> *(added by authors);* actions mainly taken at the national level are in normal type

The CHX strategy envisaged distribution through three delivery channels that correspond to the locations in which women give birth, namely public facilities, private facilities, and community/home. It identified ways to leverage existing systems, processes, and markets to ensure gains in coverage would be sustained. Within Nigeria’s decentralized health system, operationalizing this national plan required implementation, contextualized operational planning, and implementation by each of Nigeria’s 36 (plus the federal capital territory) state Ministries of Health (SMoHs) and Primary Health Care Development Agencies (SPHCDAs), which run the public primary and secondary health care facility network, as well as oversee private healthcare providers in the states. To varying degrees technical assistance partners supported individual states to develop, fund, and operationalize some or all of the state-level scale-up actions. Figure 2 summarizes key milestones in the timeline of CHX introduction and scale-up.

**Figure 2:**
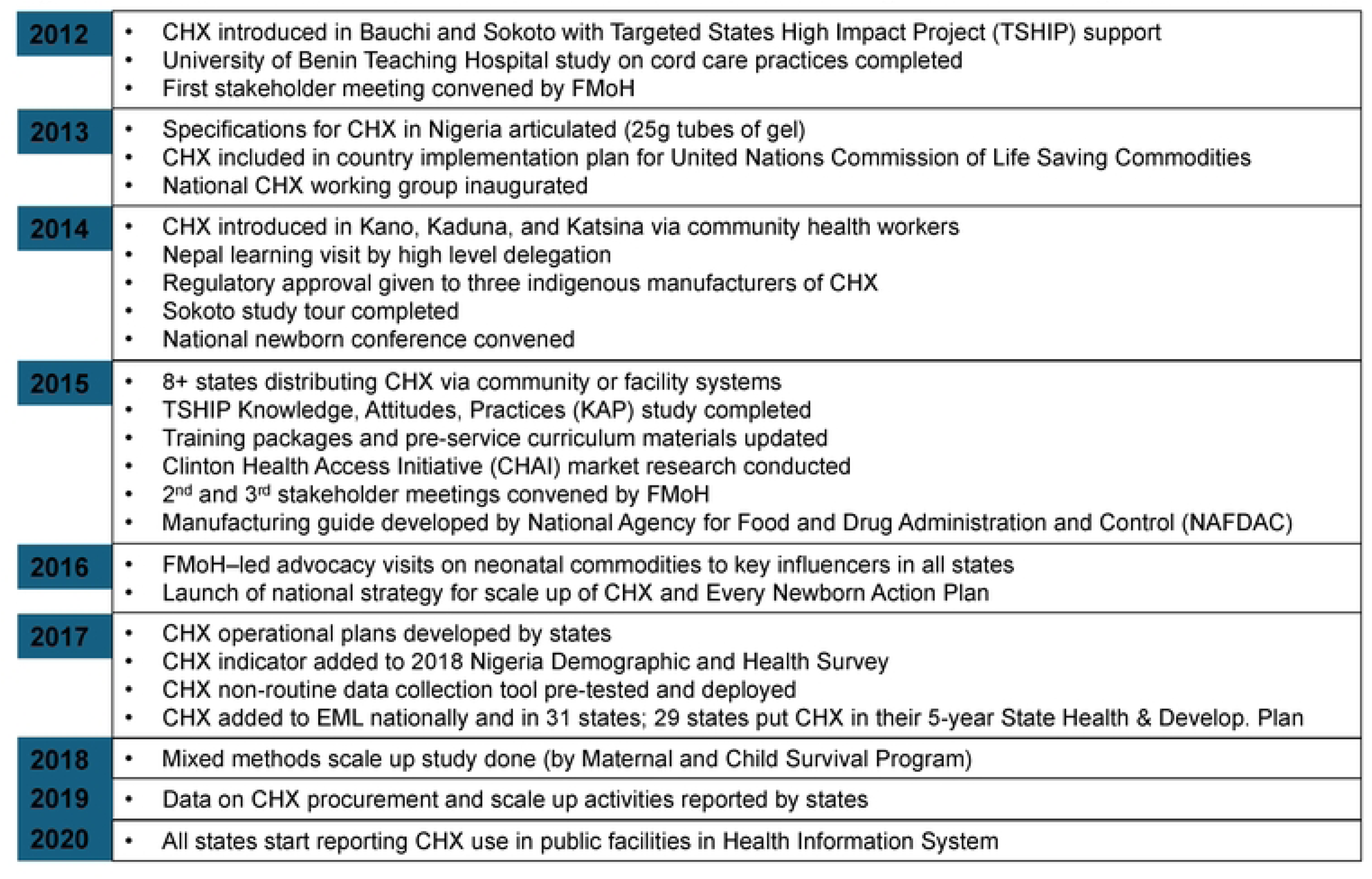
**Key milestones in the introduction and scale-up of CHX in Nigeria [based on figure in 26]**

Nigeria has continued supporting CHX scale-up, with the development of a follow-on five-year national scale-up plan that was approved in 2023 [27].

## MATERIALS AND METHODS

This was a mixed methods case study carried out longitudinally, starting with initial data collection in 2018 at the midline and going beyond the endline of the national strategy period in 2021. This study builds on and extends work that was summarized in a mixed methods case study report disseminated in 2020 that assessed progress and challenges that Nigeria experienced with the national scale-up up to that time. The team benchmarked its conduct of this longitudinal study against the Standards for Reporting Qualitative Research (SRQR) [28].

### Reflexivity statement

The study team varied over time; however, its core has remained constant and is composed of people with first-hand knowledge of CHX implementation-related events from 2012 to the present who currently or previously worked for the FMoH Child Health Division or allied international organizations that supported introduction and/or scale-up of CHX. The team reviewed critical data and documents that describe plans or assess progress at midline and endline [26,27]; convened focus groups to gather information on institutionalization and implementation of national scale-up actions; carried out key informant interviews to gather more in-depth information. The focus groups included some of the authors plus other stakeholders knowledgeable about the state of CHX scale-up in the country. This group also administered two surveys of State Reproductive Health Coordinators (SRHCs) at midline (2018) and after the endline of the national plan (2023). The fact that all study team members were involved in the scale-up process that was being studied ran the risk of bias, but this was minimized by using standard tools for measurement with explicit benchmarks and always involving the whole study team for consensus on the analysis of key findings.

### Study tools and procedures

#### Key informant interviews

A broad group of 40 key informants at the national level and in three purposively selected states were interviewed to deepen insights about the level of institutionalization and the implementation of scale-up actions mandated in the national plan. This is described in detail elsewhere [22]. The states were chosen as exemplars of locations with high levels births in public facilities (Kogi); high levels of births in private facilities (Ogun); and high levels of birth in the community (Sokoto).

#### Focus groups of national experts

In order to assess the extent of institutionalization of CHX use in the health systems the study team used a tool developed and used for other country scale-up case studies as described elsewhere [29]. The study team used this tool during facilitated group discussions with five well-informed experts in 2018 and seven in 2023. These senior experts were from FMoH, professional societies, and donor/technical agencies. The tool assesses the extent to which CHX has become integrated in systems and part of routine practice, based on assessing integration into the six health system building blocks outlined by the World Health Organization (WHO) [18]. To the usual six building blocks of governance, finance, human resources, information, supply, and service delivery the team added a seventh (demand creation) since this was a key piece of Nigeria’s national plan. In order to facilitate understanding and measurement, the team sub-divided several of the conceptually more complex building blocks, resulting in a total of 12 granular sub-components, shown in Table 1. For instance, the building block of governance was sub-divided into policy, planning, coordination, and leadership.

**Table 1:**
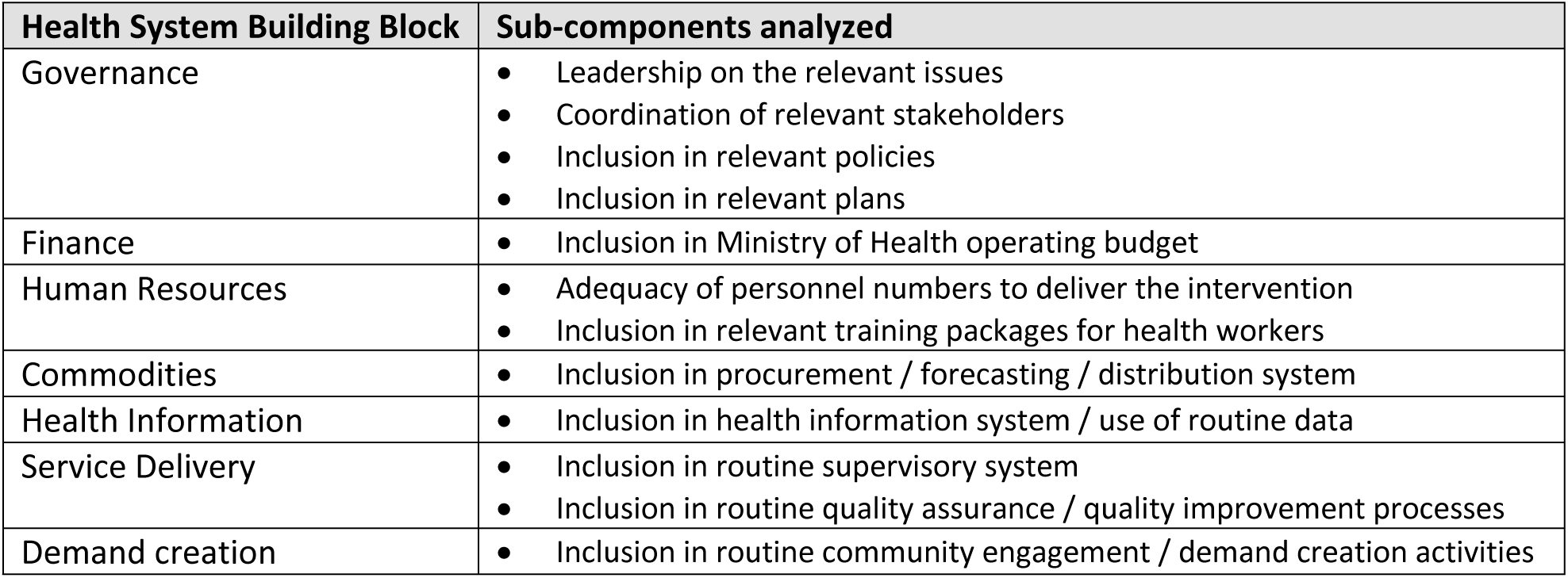
WHO health system building blocks and sub-components assessed in institutionalization analysis.

| Health System Building Block | Sub-components analyzed |
| --- | --- |
| Governance | • Leadership on the relevant issues<br>• Coordination of relevant stakeholders<br>• Inclusion in relevant policies<br>• Inclusion in relevant plans |
| Finance | • Inclusion in Ministry of Health operating budget |
| Human Resources | • Adequacy of personnel numbers to deliver the intervention<br>• Inclusion in relevant training packages for health workers |
| Commodities | • Inclusion in procurement / forecasting / distribution system |
| Health Information | • Inclusion in health information system / use of routine data |
| Service Delivery | • Inclusion in routine supervisory system<br>• Inclusion in routine quality assurance / quality improvement processes |
| Demand creation | • Inclusion in routine community engagement / demand creation activities |

For each sub-component a score of one represents the piloting stage with little institutionalization or integration into routine systems such as information systems, training, etc. and a score of four indicates complete institutionalization. In a 90-minute facilitated discussion, the experts were introduced to the definition of each sub-component, the scoring scale, and the meaning of each level in the scale. After a short discussion, the group was asked to come to a consensus on the score for institutionalization at the national level for each sub-component and provide justification for that score. A study team member facilitated the 2018 discussion, representing the ‘midline’ of the national strategy period. This group was also asked to retrospectively score the national level status for each sub-component for the year before the national strategy was issued (e.g. 2015), representing the ‘baseline’. In 2023, the study team again gathered a group of seven senior experts, four of whom had participated in the 2018 exercise, to repeat the institutionalization analysis by retrospectively considering the situation at the ‘endline’ of the 2016-2021 national strategy. The tool, scores, and justification for the scores are shown in Supplementary Materials Appendix 1.

The 2016 national strategy shown in Figure 1 envisaged that ten scale-up actions would occur at the national level. Some actions from the national strategy were disaggregated for clarity when they were discussed. Actions were scored as ‘completed’, ‘partially done’, or ‘not done.’ To assess to what extent these actions were implemented, the study team reviewed information compiled for the follow-on 2023 national strategy [27] followed by a focus-group discussion with several national level key informants in 2024 from among the authors of this paper. The results are presented in Table 3

#### Surveys of state reproductive health coordinators

Information on implementation of state-level scale-up actions was gathered in surveys sent by the FMoH to all State Reproductive Health Coordinators (SRHCs) at midline of the CHX strategy (2018) and after the endline of the strategy (2023-4). The SRHCs are responsible for and have broad knowledge of the situation for maternal, newborn, child, and reproductive health in their state. Both of these surveys asked about the implementation of state-level actions recommended in the national strategy (underlined in Figure 1) under the workstreams of “policy, advocacy, finance, and distribution”, and “marketing, user” (i.e., generating demand). Responses were obtained from all 37 SRHCs at midline (2018) and from 33 of the 37 SRHCs at endline (2023-4).

The midline SRHCs survey was less comprehensive and focused on four state scale-up actions (mobilize resources, maintain and ensure favorable policies, procure the commodity, and track progress). The endline survey asked about these plus the other four state scale-up actions in the strategy (advocate, disseminate key messages, train health workers, and generate demand). Within the domains of the state level scale-up actions, there were some adjustments to the questions from the midline to the endline surveys, based on the issues that were most relevant at the time of the survey. For instance, in 2018 an indicator for use of CHX had not yet been included in the national HIS but was being tracked in some states through a parallel non-routine data collection tool that FMoH had approved but only some states had put into implementation. So, the 2018 survey asked about use of this data collection tool. By endline the CHX use indicator had been included in the HIS, so the 2023-4 survey no longer asked about special data collection but instead asked about the completeness and timeliness of reporting of this indicator within the HIS. The surveys asked if scale-up actions had been implemented, with binary responses (Yes/No), with space for explanations. The survey tools are shown in Supplementary Materials Appendix 2. The responses from SRH Coordinators were validated by consulting the state directors of the Primary Health Care Development Agency (PHCDA). In the few cases where the two reports disagreed, the response from the state director was used. In cases where both were unsure of the answer, the response was left blank.

**Table 2:**
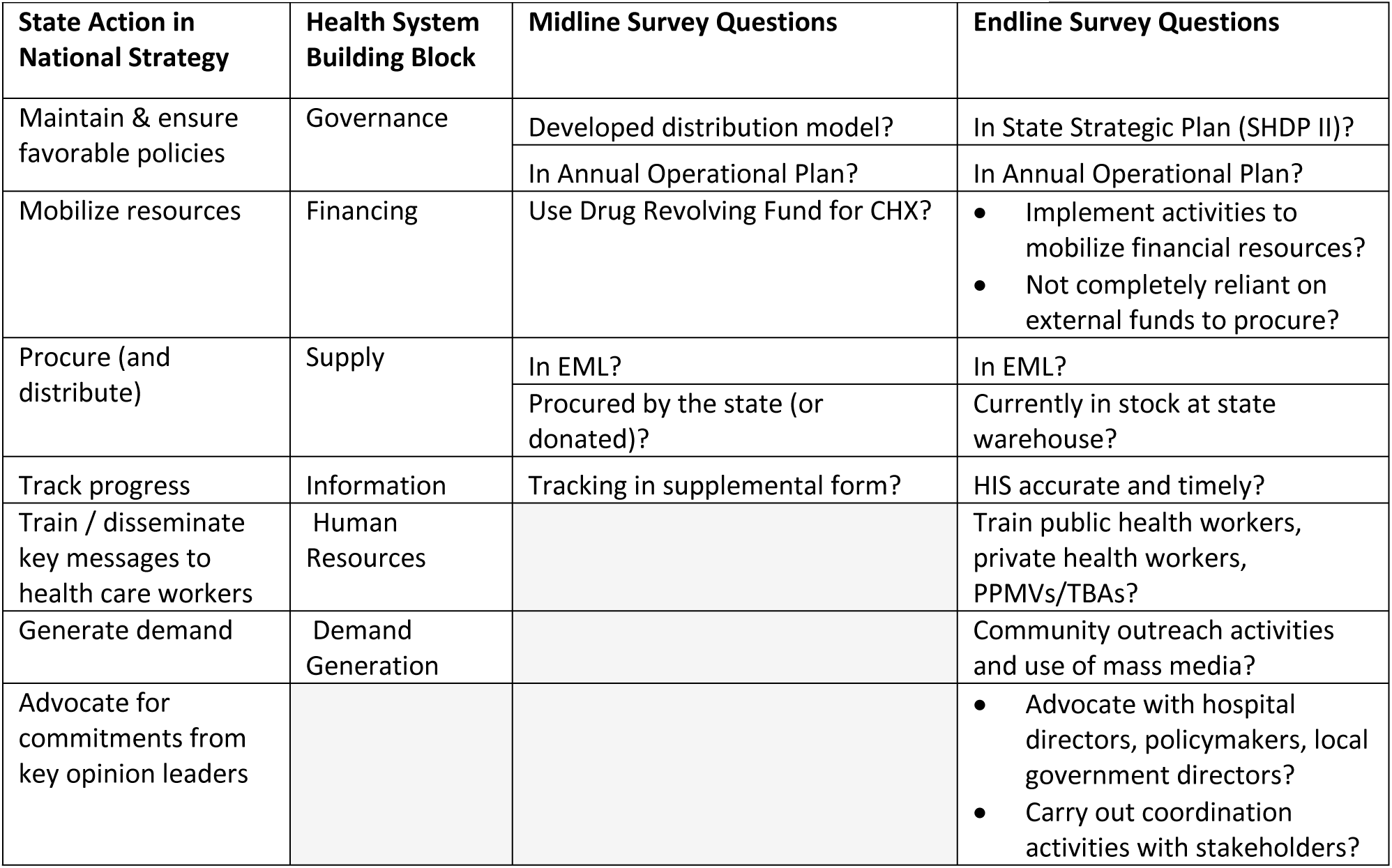
Questions in state reproductive health coordinator surveys, corresponding actions in National Strategy, categorized by WHO health system building blocks.

| State Action in National Strategy | Health System Building Block | Midline Survey Questions | Endline Survey Questions |
| --- | --- | --- | --- |
| Maintain & ensure favorable policies | Governance | Developed distribution model? | In State Strategic Plan (SHDP II)? |
|  |  | In Annual Operational Plan? | In Annual Operational Plan? |
| Mobilize resources | Financing | Use Drug Revolving Fund for CHX? | Implement activities to mobilize financial resources?<br>Not completely reliant on external funds to procure? |
| Procure (and distribute) | Supply | In EML? | In EML? |
|  |  | Procured by the state (or donated)? | Currently in stock at state warehouse? |
| Track progress | Information | Tracking in supplemental form? | HIS accurate and timely? |
| Train / disseminate key messages to health care workers | Human Resources |  | Train public health workers, private health workers, PPMVs/TBAs? |
| Generate demand | Demand Generation |  | Community outreach activities and use of mass media? |
| Advocate for commitments from key opinion leaders |  |  | Advocate with hospital directors, policymakers, local government directors?<br>Carry out coordination activities with stakeholders? |

### Ethical considerations

The Nigeria National Health Research Ethics Committee provided ethical approval and the Johns Hopkins Bloomberg School of Public Health Institutional Review Board determined that this was Non-Human Subjects Research.

## RESULTS

At the national level, institutionalization of CHX steadily progressed across all health system building blocks from baseline in 2015 to midline in 201to endline in 2021, as shown in Figure 3. Institutionalization progressed most rapidly in the health system building block of governance (i.e., policy, planning, coordination, leadership); followed by the building blocks more proximal to service delivery (i.e., human resources (personnel/training), commodities, health information). The building block that has lagged the most has been financing. Results below are discussed in reference to the six building blocks

**Figure 3:**
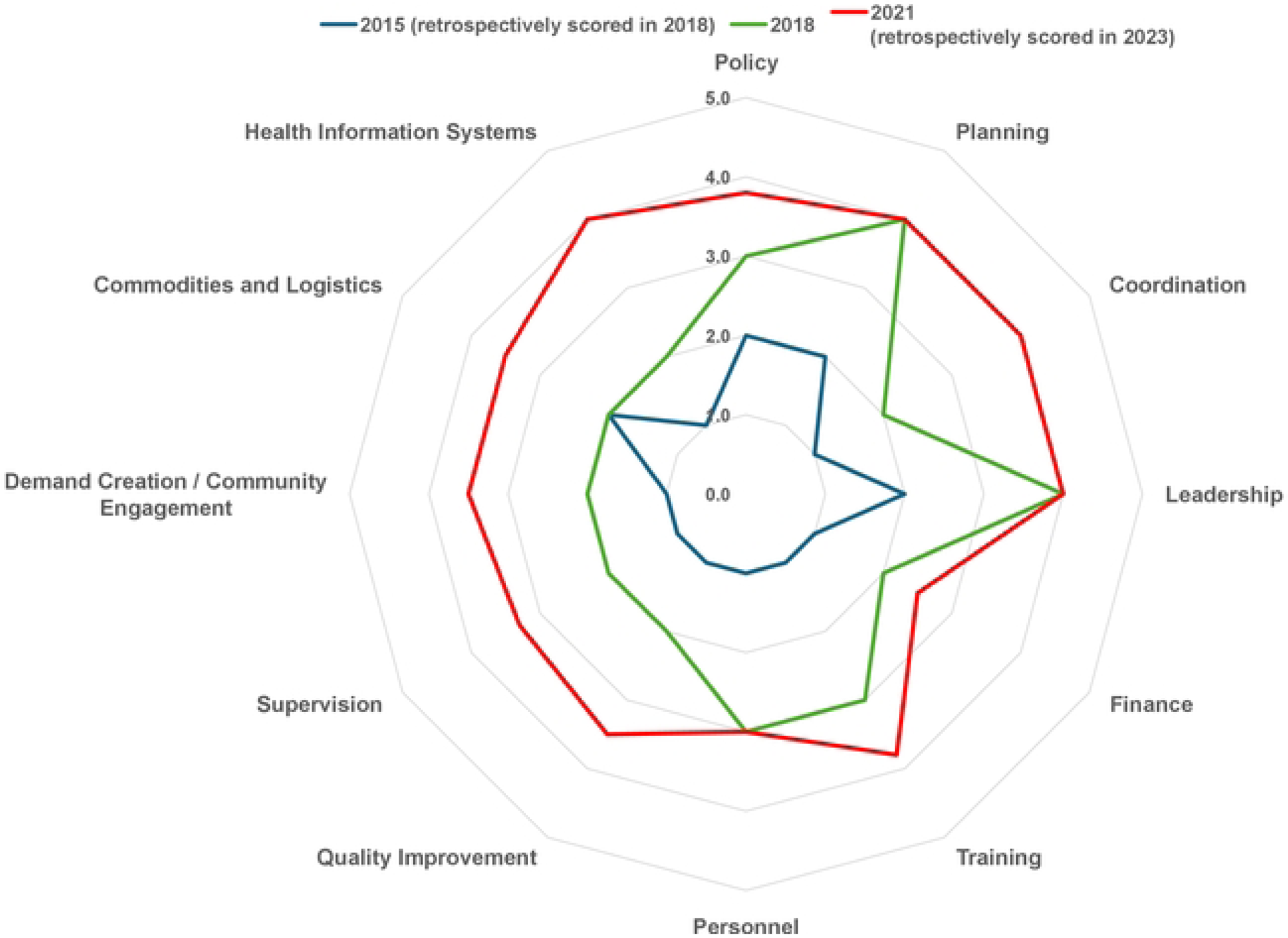
**Evolution of institutionalization of CHX in routine health systems, 2015 - 2021**

### Early institutionalization: governance

The first components to be institutionalized nationally were various aspects of governance including CHX policy formation, leadership, planning, and coordination. The 2016 national strategy included a timeline and cost analysis; however, it was still at a high level and needed to be fleshed out and contextualized by each state. CHX was put on the national Essential Medicines List (EML) in 2016. By the midline, 31 states had included CHX on their state-level EML. Although the experts in the focus group rated coordination highly, this task has been made more difficult by the fact that international partners tend to work with a small number of states and do not always share development priorities. US government-funded maternal, newborn and child health (MNCH) programs during the early in the period of the national strategy were focused on the states of Kogi and Ebonyi through the Maternal and Child Survival Program [30], then from 2019 onward in Bauchi, Ebonyi, Kebbi, Sokoto, and the FCT (Abuja) through the Integrated Health Program [31]. Although Unicef had a presence in all states during this period, they focused non-emergency MNCH programming with UK funds in in the five northern states [32] of Jigawa, Katsina, Kebbi, Yobe, and Zamfara; and through MNCH Weeks [32], they supported several other states (Benue, Ebonyi, Akwa Ibom, Ekiti, FCT).

### Later components of institutionalization: service delivery-related

System components that took longer to institutionalize were those associated with front-line implementation including training of health workers, supply chain management, and incorporation of key indicators within the HIS. Domestic production of CHX facilitated availability and improvements in supply. This was, in turn, aided by the fact that the Nigerian regulatory body prohibited importing CHX from other countries [22]. The Government of Nigeria helped pave the way by introducing a separate, expedited process for registering locally-produced essential health commodities such as CHX and abolishing import duties for raw pharmaceutical ingredients. It also specifically created a manufacturing guide for CHX, and designated CHX as an over-the-counter drug to broaden availability in the private market [22]. The absence of a routine HIS indicator led to advocacy for an officially sanctioned supplementary data-collection mechanism, which occurred in 2017, but not all states took up use of this form immediately [22]. When the FMoH updated its registers in 2020, they included CHX use and, by 2021, reporting was taking place across all states, as confirmed by the study team’s review of national level HIS reporting. Uptake of CHX was further complicated by negative publicity from a couple highly publicized tragedies in which imported CHX in packages resembling eye drops were mistakenly applied to the eyes of newborns in Adamawa and Yobe [34,35] causing blindness.

### Lagging component of institutionalization: Financing

From 2016 to 2021, the World Bank contributed financing to the states’ health and development plans through the Saving One Million Lives (SOML) initiative [36] whereby if a state included CHX in its Annual Operational Plan SOML financing could contribute toward implementation. Because of competing priorities, only 29 states did so, and even among the states that utilized SOML funds, there were large disparities in performance. Consequently, only a minority of states received additional performance-based funding [36]. Since the SOML program ended in 2021, it has become even more important that states dedicate domestic financing for continued maintenance and expansion of CHX implementation.

#### National-level scale-up actions

In terms of implementation of the ten national-level scale-up actions outlined in the 2016 strategy, Table 3 shows the consensus of the expert group. By the endline, seven of the ten actions were carried out as planned. Among these were the establishment of a newborn sub-committee as a sustainable national coordination mechanism. This committee appointed a National Uptake Coordinator in 2016 who worked in a secondment to the FMoH through 2019, using external funding, to help ensure that the scale-up process unfolded effectively [22]. Formative research was done to develop a low-literacy package insert [22]. An initial procurement was done for all states. Initial rollout was overseen in 2015, with a tailored approach in which the focus CHX distribution channel in each state was predicated on the predominant location of births. Policies were updated as needed. In addition to tracking use in the HIS, packaging of CHX in bottles similar to those used for eye drops was banned after the 2015 incidents in Yobe and Adamawa mentioned above [22].

The other three national actions were only partially carried out: supporting local manufacturers, establishing and strengthening state-level coordination, and monitoring evidence from further studies. Nigerian manufacturers (initially four and later three) were incentivized to supply CHX but registration of the product was slow, causing an impasse at midline of the national plan as the government of Nigeria had stopped issuing import waivers in order to spur local production. Complicating matters, some organizations, notably Unicef, were statutorily unable to purchase from local manufacturers without WHO pre-qualification, which was slow in coming [22]. The mismatch between supply and demand has been an ongoing issue, as manufacturers continue to say that they have the capacity to supply enough CHX for complete national population coverage, but that states and private entities have not matched this potential capacity with sufficient demand [27]. There was also insufficient coordination at the state level, where coordination and tracking progress on CHX implementation became one of many responsibilities of already over-stretched State RH Coordinators [25]. Monitoring the literature for new CHX studies has been sporadic, as the Newborn Sub-Committee of the Child Health Technical Working Group – which was considered the most appropriate federal-level coordination mechanism – has other competing responsibilities.

**Table 3:** Implementation of national level actions envisaged in national plan. Green = completely implemented; yellow = partially implemented

| National level action in 2016 national strategy | Achievement |
| --- | --- |
| Improve product packaging | Complete |
| Monitor evidence from future studies | Partial |
| Appoint uptake coordinator | Complete |
| Formalize national coordination mechanism | Complete |
| Establish and strengthen coordination in each state | Partial |
| Oversee strategic approach to rollout | Complete |
| Leverage existing public and private channels | Complete |
| Support local manufacturers to expand distribution | Partial |
| Initial procurement | Complete |
| Maintain favorable policies | Complete |

#### Implementation of scale-up actions at the state level

Table 4 shows the number and percent of states that had implemented each of the state-level scale-up actions envisaged in the national plan by midline and endline. The full results of the surveys are in Supplementary Materials Appendix 3. Only the endline survey asked about training, demand generation, and advocacy. There was wide variability in terms of the number of actions implemented by states. At midline there were several states that had not implemented any of the mandated actions (see Appendix 3). Whereas 11 (30%) had implemented 5 of the 6 mandated key actions, but none had implemented all six. The greatest number of states put CHX on their state EML. At midline the only actions implemented by fewer than half of states were financing ongoing procurement by putting CHX in their state Drug Revolving Fund and tracking CHX in the FMoH-mandated supplemental register. At endline, there were four states that did not respond to the survey and some questions on completed surveys left blank. The actions implemented the least were inclusion in the State Health and Development Plan (22 of 33 (36%)), mobilization of funds (8 of 33 (24%)), and maintenance of stock in state warehouse (11 of 29 (24%)).

**Table 4:** Number (and %) of states that implemented state-level scale-up actions at midline and endline.

| MIDLINE state-level scale-up actions | No. states implement / No. states with responses | % states implementing action | ENDLINE state-level scale-up actions | No. states implement / No. states with responses | % states implementing action |
| --- | --- | --- | --- | --- | --- |
| State developed CHX distribution model | 21 / 37 | 57% | CHX in State Health and Development Plan | 12 / 33 | 36% |
| CHX on Annual Operational Plan | 25 / 37 | 68% | CHX on Annual Operational Plan | 22 / 33 | 67% |
| CHX in Drug Revolving Fund | 13 / 37 | 35% | Mobilize funds for CHX program<br>Not completely reliant on external funding for CHX program | 8 / 33<br>15 / 31 | 24%<br>48% |
| CHX placed on state EML | 23 / 37 | 62% | CHX placed on state EML | 27 / 33 | 82% |
| CHX procured by state | 23 / 37 | 62% | CHX in stock at state warehouse | 11 / 29 | 24%<br>48% |
| CHX tracked in supplemental register | 15 / 37 | 41% | CHX regularly tracked in HIS<br>CHX information in HIS is accurate | 37 / 37<br>19 / 25 | 100%<br>76% |
|  |  |  | Train health workers on use of CHX | 26 / 33 | 79% |
|  |  |  | Demand generation activities for CHX | 29 / 33 | 88% |
|  |  |  | Advocate with decisionmakers on use of CHX<br>Coordinate stakeholders for CHX programming | 15 / 33<br>16 / 33 | 45%<br>48% |

\* From review of HIS by the study team. All states had the indicator in their registers and data for all 12 monthly reports in 2021.

## DISCUSSION

A widely cited paper in the field of translational research [37] concluded that it takes an average of 17 years for an innovation to achieve widespread impact at scale in developed countries. These authors said that their investigation was hampered by the lack of clear definitions about “starting points” and “end points.” In the case of Nigeria, a reasonable starting point would be when the TSHIP project initiated the first introduction of CHX in the country in 2012. By this reckoning, after only nine years Nigeria had achieved substantial progress in terms of institutionalizing the intervention in routine systems by the end of its initial national scale-up plan, in 2021. While well on its way to full large-scale system integration of CHX, the work is not complete.

Many who have studied scale-up point out that the task is iterative in nature and does not typically follow a straight line because of shifting priorities of implementers, donors and technical agencies as well as unforeseeable changes in the larger context [38,39]. For example, the global COVID pandemic caused severe interruptions to routine MNCH services in Nigeria from early 2020 until the end of the national plan period in 2021, in terms of both service disruption and the need to re-orient health system priorities to respond to the emergency [40,41,42].

### Institutionalization in health system building blocks, including sequencing of the process

Across the health system building blocks, Nigerian decisionmakers followed a sequence for institutionalizing CHX that made sense from a practical perspective, and usually (but not always) also made sense from a technical perspective. First, policymakers developed the basic ‘rules of the game’ through governance mechanisms – policies, leadership, planning, and coordination structures – while contemporaneously encouraging a ramp-up of domestic CHX production. Later they developed operational support mechanisms to aid implementation of the plans. These included training, logistics, and monitoring mechanisms.

There are two instances where technical and practical sequencing considerations diverged. One is the development of domestic financing, which has been the slowest component to develop. This lag in domestic financing is unsurprising, given the large number of competing priorities and the scarcity of resources to meet those needs; however, this has left progress vulnerable. As international donor funding priorities inevitably shifted and as the COVID pandemic intervened, a number of states decreased their attention to the continuing needs of the CHX scale-up effort. Another example is the timeline for revision of the health information system to include the CHX indicator. The indicator was not added until the fourth year of the national scale-up strategy, even though it would have been logical for it to have occurred much earlier so that decisionmakers could have frequent and universal information on progress. But as in many countries, registers are infrequently revised in Nigeria due to cost and logistical difficulties, so the inclusion of CHX in registers needed to wait until the next scheduled revision. Although a number of states used an FMoH-mandated supplemental register for CHX, a significant number did not and therefore were ‘driving blind’ without critical information needed to actively manage the scale-up process.

There have been ongoing discussions within the FMoH about the desire to rapidly integrate CHX into the health system (personal communication, Director of Maternal Child Health, Nigeria FMoH). This consideration led to the elimination by midline of the National Uptake Coordinator position that was specifically devoted to CHX scale-up and shift the task of coordination of the CHX scale-up process to the established Newborn Sub-Committee of the Child Health Technical Working Group. This shift may have been premature because the process seems to have lost momentum, as the Newborn Sub-Committee has a wide array of competing priorities. It makes sense that once institutionalized (i.e., integrated into routine systems), there should be no separate effort on, for instance, data collection or health worker training specifically for CHX. But institutionalization is not an all-or-none proposition, and there are some health systems building blocks, like finance, that still need separate and concerted effort if Nigeria is to continue making progress in its CHX scaling journey. This is the sort of issue that a dedicated national coordinator, working in concert with a scale-up management team, could help address.

Another consideration is that once CHX has been fully institutionalized in a health system building block, it is only reasonable to expect that the system will work as well for CHX as it does for other interventions that depend on that system. An example is the logistics system, where stockouts have continued to hamper the efforts to deliver CHX in some states. This is not a reflection of a deficiency in the scale-up process for CHX, but rather a reflection of wider health systems deficiencies that require a broader response.

### Implementation of actions to support scale-up

#### National level

The development of the 2016 national strategy was clearly catalytic for scale-up of CHX. The FMoH took the experience of other countries, distilled and contextualized it, and developed a detailed action plan for the national level as well as a template for actions by states. The latter was a critical step for Nigeria’s highly decentralized system and provided an impetus for progress in every state. The fact that the FMoH developed a follow-up CHX national scale-up plan in 2024 is indicative of the fact that scale-up of CHX is a locally-driven initiative, even while it has been intermittently supported by different international partners, in various state locations, and in diverse ways. Even so, the new national scale-up strategy acknowledges weaknesses in coordination, which is a requirement for navigating the complexity of Nigeria’s decentralized health system [27]. Another key aspect of the scale-up was the establishment of in-country capability to manufacture and distribute the volume of CHX needed to cover 100% of country need, but there have been ongoing growing pains in matching potential supply with sufficient demand [27].

#### State level

It is striking how quickly certain scale-up actions were implemented and governance functions were institutionalized across the national and state levels, even in Nigeria’s highly decentralized system. By midline of the national strategy a majority of states made initial procurements, had included CHX on their state EML, and included it in their annual operational plan. The slowing of effort can be seen in the endline assessment. Only a few of the remaining states had added CHX to their EML, and no more states than at midline had included it in their annual operational plan.

### Study Limitations

The state coordinators who answered the surveys may have had incomplete knowledge of the scale-up actions taken in their state. However, almost all had been at their job for over a year, and their responses were validated by the state director of the PHCDA. Between the two respondents, in all cases at least one had been in their job for longer than a year, and most for much longer. The SRHC surveys also did not determine the level of intensity of implemented actions, but only asked for a binary (Yes/No) response. Finally, the endline survey was done about two years after the end of the national strategy. This could have caused misclassification of the state of implementation of the state-level scale-up actions, as respondents’ memories might have been imperfect.

## CONCLUSIONS

This case study of Nigeria’s ongoing journey in scaling up CHX provides important lessons on the value of systematically developing a clear and explicit scale-up strategy for expanding and institutionalizing a new intervention in a highly decentralized system, especially one that includes a public-private partnership, in this case with drug manufacturers. Even with the complex interplay between specific contextual and programmatic factors, this analysis offers generalizable lessons for making scale-up strategies more effective. First, it highlights the need for sustained action over 10 or more years, a much longer time horizon than any one project lasts. In such a prolonged and complex environment, the often-used metaphor in the popular mind and among some that study scale-up of an intervention package that flows in an ever-spreading stream is a poor one. Instead, if successful there will be multiple streams of action that need to mesh and contribute to the expansion of an intervention package (i.e., through the action of multiple projects, various actors, and overlapping strategies). All these will “flow together” and contribute to the gathering momentum in a landscape that is very likely to be shifting. Second, the scale-up process required coordinated action across several key public and private organizations. The National Uptake Coordinator aided in that coordination across organizations and over time, but unfortunately, the fact that the position was donor-funded meant that it disappeared prematurely while the need for continued coordination remained. So then an overstretched Newborn Sub-Committee had difficulty continuing with the intensity of action needed to maintain progress. Third, this case study highlights the fact that steps towards institutionalizing key components required for successful scale-up can be constrained by practical considerations (like when data collection registers are updated) and unknowable developments (like the COVID pandemic). Such ‘real-world’ considerations can make the scale-up process much more difficult. These are the sort of constraints that are not amenable to testing through conventional rigorous study designs like randomized controlled trials. This makes the evidence gathering for successful strategies exceedingly difficult. Lastly, this analysis demonstrates that as institutionalization progresses, the scaled intervention can only be as strong as the larger health system into which it is integrated, as illustrated by ongoing challenges with the accuracy of monitoring information, general financing constraints and difficulties with commodity stockouts. These are the sorts of weaknesses that are compensated for in smaller ‘proof of concept’ trials, but cannot be avoided in the ‘real-world’ of a national scale-up experience. Taken together, all these lessons point to the need for a coalition of stakeholders undertaking a long-term and active management of the scale-up process that adapts to unforeseeable changes. Future action for scale-up of CHX in Nigeria should be directed towards robust state-level activities to ensure steady financing, improve commodity logistics, ensure coordination and improve monitoring. Nigeria’s follow-on five-year strategy wisely includes these elements.

## Data Availability

All underlying data is contained in the Supplementary Materials (Appendices 1,2, and 3).

## ACKNOWLEDGEMENTS

The authors would like to acknowledge the significant contribution of Suleiman Yakubu who led the data collection for the key informant interviews in 2019. We want to thank Ruth Simmons and Peter Fajans of the ExpandNet Secretariat who gave advice during the implementation process and reviewed early drafts of the manuscript. We would also like to acknowledge Stephen Hodgins of the University of Alberta and Larry Cooley of MSI International who advised the group studying scale up during the period of the national strategy. We want to acknowledge Anuja Shah, Jennifer Hoeg, Susan Gigli, and Jenna Wright, who contributed to the effort to support and study scale-up. Finally, we want to acknowledge the heroic and ongoing efforts of health providers across Nigeria who strive to reduce newborn mortality through the use of CHX and other means.

## APPENDIX 1 SUPPLEMENTAL MATERIAL

State Reproductive Health Coordinator Surveys

## APPENDIX 2 SUPPLEMENTAL MATERIAL

Institutionalization Scoring Matrix

## APPENDIX 3 SUPPLEMENTAL MATERIAL

State Reproductive Health Coordinator Survey Responses

